# Sociodemographic and socioeconomic disparities in COVID-19 vaccine uptake in Belgium – A nationwide record linkage study

**DOI:** 10.1101/2023.01.31.23285233

**Authors:** Lisa Cavillot, Joris A.F van Loenhout, Brecht Devleesschauwer, Chloé Wyndham-Thomas, Herman Van Oyen, Jinane Ghattas, Koen Blot, Laura Van den Borre, Matthieu Billuart, Niko Speybroeck, Robby De Pauw, Veerle Stouten, Lucy Catteau, Pierre Hubin

**Author notes:** Correspondence **Lisa Cavillot**, Department of Epidemiology and Public Health, Sciensano, Rue Ernest Blerot 1, 1070 Anderlecht, Belgique.

## Abstract

**Background:** Recent studies have identified important social inequalities in SARS-CoV-2 infection and related COVID-19 outcomes in the Belgian population. The aim of our study was to investigate the sociodemographic and socioeconomic characteristics associated with the uptake of COVID-19 vaccine in Belgium.

**Methods:** We conducted a cross-sectional analysis of the uptake of a first COVID-19 vaccine dose among 5,342,110 adults (≥18 years) in Belgium from December 28^th^ 2020 (official starting date of the vaccination campaign) until August 31^st^ 2021. We integrated data from four national data sources: the Belgian vaccine register (vaccination status), COVID-19 Healthdata (laboratory test results), DEMOBEL (sociodemographic/socioeconomic data), and the Common Base Registry for HealthCare Actors (individuals licensed to practice a healthcare profession in Belgium). We used multivariable logistic regression analysis for identifying characteristics associated with not having obtained a first COVID-19 vaccine dose in Belgium and for each of its three regions (Flanders, Brussels, and Wallonia).

**Results:** During the study period, 10% (536,716/5,342,110) of the Belgian adult population included in our study sample was not vaccinated with a first COVID-19 vaccine dose. A lower COVID-19 vaccine uptake was found among young individuals, men, migrants, single parents, one-person households, and disadvantaged socioeconomic groups (with lower levels of income and education, unemployed). Overall, the sociodemographic and socioeconomic disparities were comparable for all regions.

**Conclusions:** The identification of sociodemographic and socioeconomic disparities in COVID-19 vaccination uptake is critical to develop strategies guaranteeing a more equitable vaccination coverage of the Belgian adult population.

## Background

The presence of a social gradient in the Severe Acute Respiratory Syndrome Coronavirus 2 (SARS-CoV-2) infections and subsequent coronavirus disease 2019 (COVID-19) outcomes has been clearly demonstrated through various studies in different regions of the world [1]. Certain sociodemographic (SD) groups, such as the elderly, men, and individuals from ethnic minorities, as well as individuals from disadvantaged socioeconomic (SE) groups, have shown higher risks of getting infected by SARS-CoV-2, developing severe complications after infection (e.g. hospitalization, intensive care unit admission, death), and suffering from indirect complications (e.g. education and employment losses, housing instability, food scarcity) [2,3]. In Belgium, the same patterns have been identified, whereby the most disadvantaged SE groups have a higher incidence of SARS-CoV-2 infection and presented higher levels of excess mortality during the COVID-19 pandemic [4–6].

Vaccination has been established as a powerful public health instrument to decrease transmission of the virus and lower the probability of developing severe COVID-19 health outcomes [7,8]. Despite widespread vaccine promotion efforts, concerns regarding vaccine equity remain. Although higher COVID-19 incidence and mortality have been described for SE disadvantaged groups, some studies have documented a lower COVID-19 vaccination coverage among them [9,10]. The vaccine uptake is influenced by a range of factors, including vaccine accessibility, awareness (information about the disease and the vaccine), and willingness which can be influenced by the socio-cultural environment, political and religious orientation, or pre-existing health needs [11–13]. Understanding the social pattern in COVID-19 vaccination uptake is crucial to improve the ongoing Belgian COVID-19 vaccination campaign. The identification of health inequalities in COVID-19 vaccination coverage may also serve for future vaccine campaigns and pandemic preparedness.

Belgium’s nationwide vaccination campaign against COVID-19 started early January 2021, after an initial pilot program in December 2020. Campaign roll-out first targeted pre-defined priority groups that were sequentially invited: nursing home residents and staff, promptly followed by hospital-based health professionals (January 2021 onwards), frontline healthcare professionals and both residents and staff of collective care facilities (February 2021 onwards), individuals aged 65 and over (in order of decreasing age), and individuals with comorbidities at increased risk of severe COVID-19 (March 2021 onwards). In June 2021, the vaccine campaign was extended to all aged 18 years and older [14,15].

The objective of the present study was to investigate whether the uptake of a first COVID-19 vaccine dose, from December 28^th^ 2020 to August 31^st^ 2021, has been equitable across Belgium and all its regions and, if not, to identify which groups of adults (≥ 18 years) have a lower COVID-19 vaccine uptake, with respect to their SD and SE characteristics.

## Methods

### Study design and data sources

In this cross-sectional study, we investigated SD and SE disparities in the uptake of a first COVID-19 vaccine dose using data from the LINK-VACC project [16]. LINK-VACC was set-up by Sciensano, the Belgian institute for health, to perform the post-authorization surveillance of COVID-19 vaccines uptake and effectiveness in Belgium. To that aim, selected variables from multiple existing national health and social sector registers were linked at an individual level using the unique Belgian social security number.

The databases were linked within a secured pseudonymised environment hosted by Healthdata.be, a data platform within Sciensano [17]. For the present study, four databases were used:

1. Vaccinnet+ the Belgian vaccine register, which contains information on COVID-19 vaccine doses administered to Belgian residents. It includes demographical data of the vaccinated person (sex, age, postal code of residence) and on the administered vaccine (brand of the vaccine, lot number, date of administration, date of registration).
2. The COVID-19 Healthdata test database containing data from COVID-19 laboratory tests performed in Belgium (including sampling data and test result) as well as demographical data on the tested person (sex, age, postal code of residence).
3. The DEMOBEL database provided by the Statistics Belgium (Statbel) which contains variables related to SD and SE characteristics as well as information on the status in the national register.
4. The Common Base Registry for HealthCare Actors (CoBRHA) allowing the identification of individuals who have been licensed to practice a healthcare profession in Belgium (by profession and specialty).

### Study population definition

The study population consists in all individuals residing in Belgium aged of 18 years and over tested at least once for COVID-19 (PCR and rapid antigen tests) in Belgium before the 31^st^ of August 2021, as recorded in the COVID-19 Healthdata test database (n=5,661,661). Thanks to an individual linkage with DEMOBEL, SD and SE data were available for 97·6% (n=5,525,634/5,661,661) of them. After this linkage, we excluded individuals who were deregistered, migrated, and deceased based on their status in the national register available in DEMOBEL database as well as individuals with either age, sex, or region unknown.

Our final study population was composed of 5,342,110 adults (≥ 18 years) tested at least once in Belgium before August 31^st^ 2021, for whom Vaccinnet+ was consulted in order to determine their vaccination status for a first COVID-19 vaccine dose. A flow chart demonstrating the process for selecting our final study population is available in Figure 1.

**Fig. 1.**
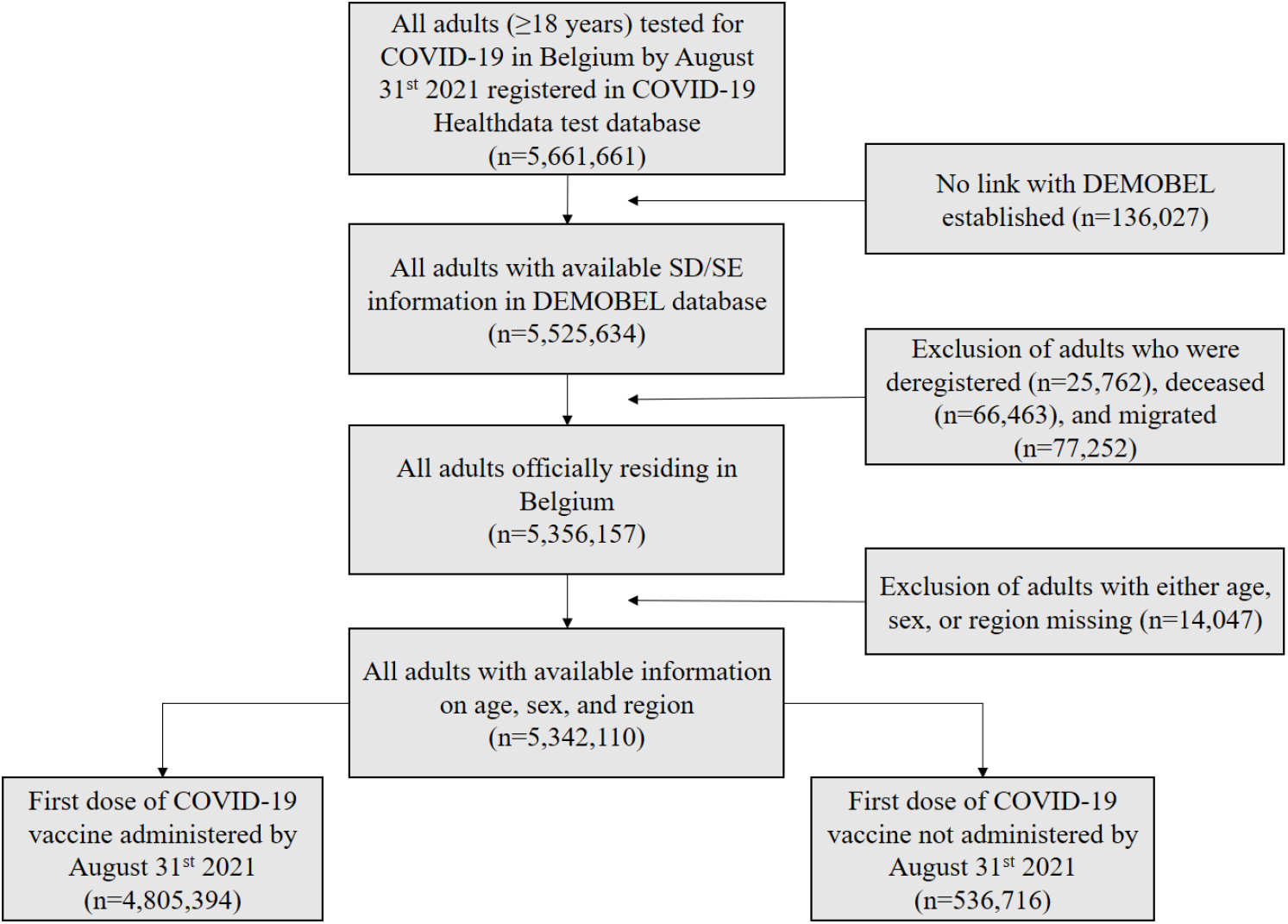
Flowchart of the study population based on the COVID-19 Healthdata test database, Belgium, December 28^th^ 2020 - August 31^st^ 2021

### Outcome

The outcome of this study was defined as a first dose of any COVID-19 vaccine (approved by the European Union) between the start of the vaccination campaign (December 28^th^ 2020) and August 31^st^ 2021. By this date, all individuals of 18 years and over officially residing in Belgium should have received an invitation to be vaccinated with a first dose and the opportunity to receive it. Figure 2 shows COVID-19 first-dose vaccination coverage over time by region and age group in Belgium. Moreover, administration of a first dose is a good predictor of a full primary course of vaccination since the proportion of fully vaccinated individuals in those who received at least one dose is close to 100% [18]. Hence, we modeled the probability for a Belgian resident not to have received a first dose of COVID-19 vaccine as of August 31^st^ 2021.

**Fig. 2.**
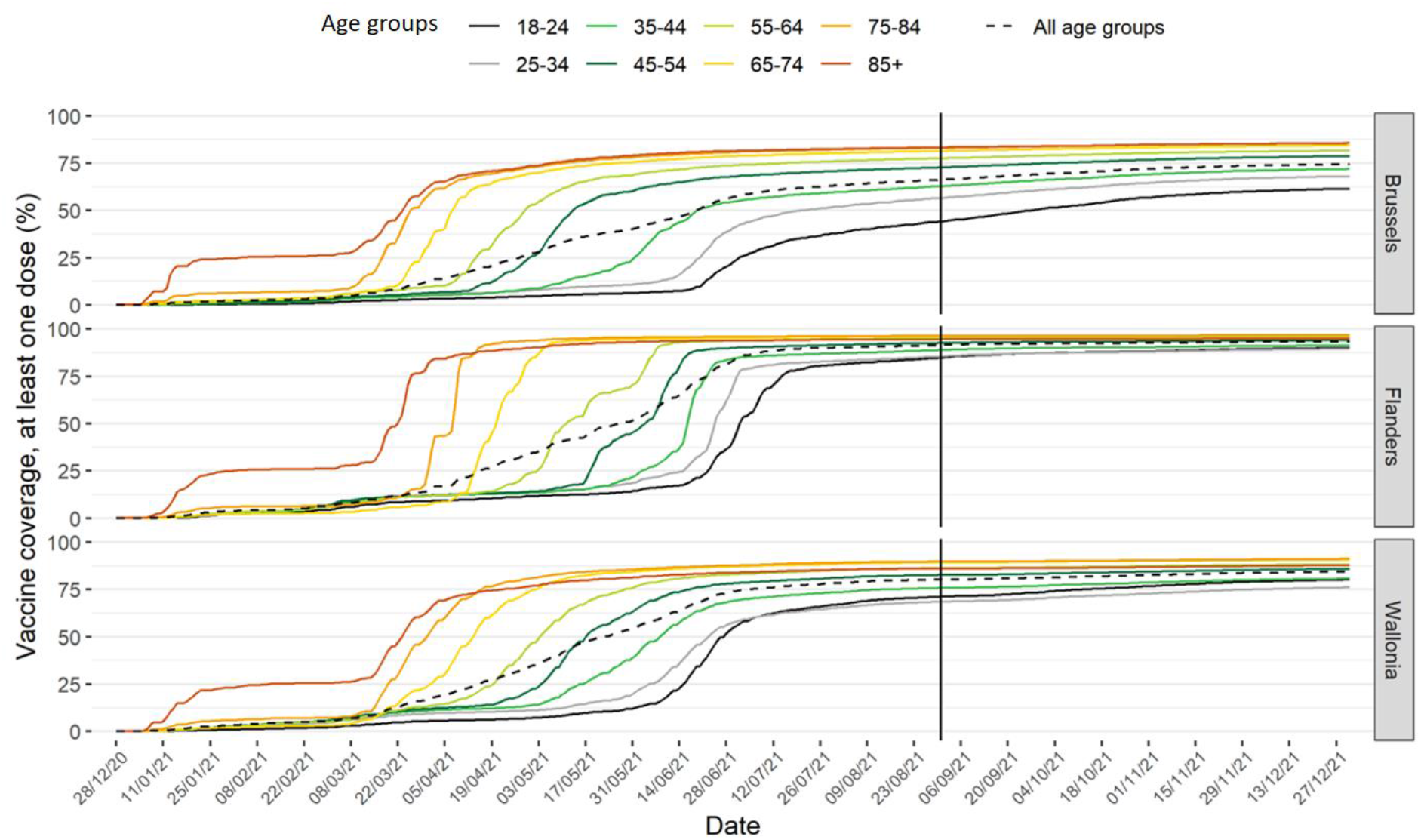
Vaccination coverage of a first dose of COVID-19 vaccine over time by age groups and regions, Belgium, December 28^th^ 2020 - December 27^th^ 2021

### Covariates

To identify characteristics associated with the uptake of a first COVID-19 vaccine dose, we focused on several relevant SD and SE characteristics (Supplementary Table 1). As SD characteristics, age, sex, region of residence, household type, and migration background were included. Age, sex, and region of residence were available in Vaccinnet+ and COVID-19 Healthdata test database, registers re-using demographic information from the Belgian national register. Household type and migration background were available in DEMOBEL database. The household type gave a partial picture of the social environment and was categorized into six groups: one-person households, couples without children, couples with children, single parents, collectivity (e.g. prison, nursing homes, religious community), other. Migration background was based on a combination of the first nationality and the parents’ country of origin. Four groups were distinguished: Belgian natives, second-generation migrants, first-generation European migrants, and first-generation non-European migrants. As SE characteristics, we included income, educational level, and employment status available in DEMOBEL. Educational level was classified in eight categories using the International Standard Classification of Education (ISCED). We merged these different categories into three main education levels: low (ISCED0 to ISCED2), moderate (ISCED3 to ISCED4), and high (ISCED5 to ISCED 8). Income information was available as deciles of the net income of the whole household. This indicator was further categorized into low income (decile 1 to 4), moderate income (decile 5 to 7), and high income (decile 8 to 10). The employment status allowed us to discern if individuals were (un)employed. Finally, we were able to determine whether individuals had a healthcare degree using a variable provided by CoBRHA. A summary table of the data holder and time frame of all SD and SE variables is available in Supplementary Table 1.

**Table 1:** Sociodemographic and socioeconomic characteristics of the population according to the uptake of a first dose of COVID-19 vaccine in Belgium, December 28^th^ 2020 - August 31^st^ 2021.

|  | All<br>(n=5,342,110 ) | First dose administered by<br>August 31<sup>st</sup> 2021<br>(n=4,805,394) | First dose not administered<br>by August 31<sup>st</sup> 2021<br>(n=536,716) |
| --- | --- | --- | --- |
| Variables | n (%<sub>column</sub>) | n (%<sub>row</sub>) | n (%<sub>row</sub>) |
| Age groups (years) |  |  |  |
| 18-24 | 645,416 (12.08) | 547,826 (84.88) | 97,590 (15.12) |
| 25-34 | 1,051,576 (19.68) | 886,846 (84.33) | 164,730 (15.67) |
| 35-44 | 978,478 (18.32) | 857,111 (87.60) | 121,367 (12.40) |
| 45-54 | 903,514 (16.91) | 827,999 (91.64) | 75,515 (8.36) |
| 55-64 | 797,786 (14.93) | 752,601 (94.34) | 45,185 (5.66) |
| 65-74 | 511,784 (9.58) | 492,092 (96.15) | 19,692 (3.85) |
| 75-84 | 296,974 (5.56) | 288,939 (97.29) | 8,035 (2.71) |
| 85+ | 156,582 (2.93) | 151,980 (97.06) | 4,602 (2.94) |
| Regions |  |  |  |
| Brussels | 579,687 (10.85) | 452,251 (78.02) | 127,436 (21.98) |
| Flemish | 3,348,547 (62.68) | 3,108,510 (92.83) | 240,037 (7.17) |
| Walloon | 1,413,876 (26.47) | 1,244,633 (88.03) | 169,243 (11.97) |
| Sex |  |  |  |
| Women | 2,849,932 (53.35) | 2,565,422 (90.02) | 284,510 (9.98) |
| Men | 2,492,178 (46.65) | 2,239,972 (89.88) | 252,206 (10.12) |
| Migration background |  |  |  |
| Belgian natives | 3,652,357 (68.37) | 3,461,899 (94.79) | 190,458 (5.21) |
| Second-generation migrants | 324,420 (6.07) | 262,465 (80.90) | 61,955 (19.10) |
| First-generation European migrants | 607,586 (11.37) | 493,062 (81.15) | 114,524 (18.85) |
| First-generation non-European migrants | 757,747 (14.18) | 587,968 (77.59) | 169,779 (22.41) |
| Household type |  |  |  |
| One-person households | 878,506 (16.44) | 786,400 (89.52) | 92,106 (10.48) |
| Collectivity | 96,213 (1.80) | 92,929 (96.59) | 3,284 (3.41) |
| Couples without children | 1,302,576 (24.38) | 1,230,134 (94.44) | 72,442 (5.56) |
| Couples with children | 2,363,467 (44.24) | 2,105,396 (89.08) | 258,071 (10.92) |
| Single parents | 548,369 (10.27) | 463,386 (84.50) | 84,983 (15.50) |
| Missing | 26,752 (0.50) | 20,574 (76.91) | 6,178 (23.09) |
| Other | 126,227 (2.36) | 106,575 (84.43) | 19,652 (15.57) |

| Variables | All<br>(n=5,342,110 ) | First dose administered by<br>August 31st 2021<br>(n=4,805,394) | First dose not administered<br>by August 31st 2021<br>(n=536,716) |
| --- | --- | --- | --- |
|  | n (%column) | n (%row) | n (%row) |
| Education level |  |  |  |
| Low | 1,567,744 (29·35) | 1,397,598 (89·15) | 170,146 (10·85) |
| Moderate | 1,569,361 (29·38) | 1,423,852 (90·73) | 145,509 (9·27) |
| High | 1,511,494 (28·29) | 1,422,694 (94·13) | 88,800 (5·87) |
| Missing | 693,511 (12·98) | 561,250 (80·93) | 132,261 (19·07) |
| Income |  |  |  |
| Low | 1,815,339 (33·98) | 1,534,829 (84·55) | 280,510 (15·45) |
| Moderate | 1,564,464 (29·29) | 1,431,971 (91·53) | 132,493 (8·47) |
| High | 1,762,607 (32·99) | 1,673,416 (94·94) | 89,191 (5·06) |
| Missing | 199,700 (3·74) | 165,178 (82·71) | 34,522 (17·29) |
| Healthcare degree |  |  |  |
| Yes | 423,852 (7·93) | 398,902 (89·59) | 24,950 (10·41) |
| No | 4,918,258 (92·07) | 4,406,492 (94·11) | 511,766 (5·89) |
| Employment status |  |  |  |
| Unemployed | 1,874,544 (35·09) | 1,695,122 (90·43) | 179,422 (9·57) |
| Employed | 3,364,795 (62·99) | 3,032,575 (90·13) | 332,220 (9·87) |
| Missing | 102,771 (1·92) | 77,697 (75·60) | 25,074 (24·40) |

n(%column): absolute number with percentage of the total population; n(%row) : absolute number with percentage of the subgroup per sociodemographic and socioeconomic characteristics.

For some individuals, there was missing information on income, household type, education level, or employment status. In such cases, those missing values were considered as a separate category as they were associated with certain SD and SE characteristics and not randomly distributed in the population (e.g. the proportion of individuals for which one of these SE characteristics is missing on the total population is about 40·0% for first-generation European migrants and first-generation non-European migrants against 5·0% for Belgian natives) (see Supplementary Table 2).

**Table 2:**
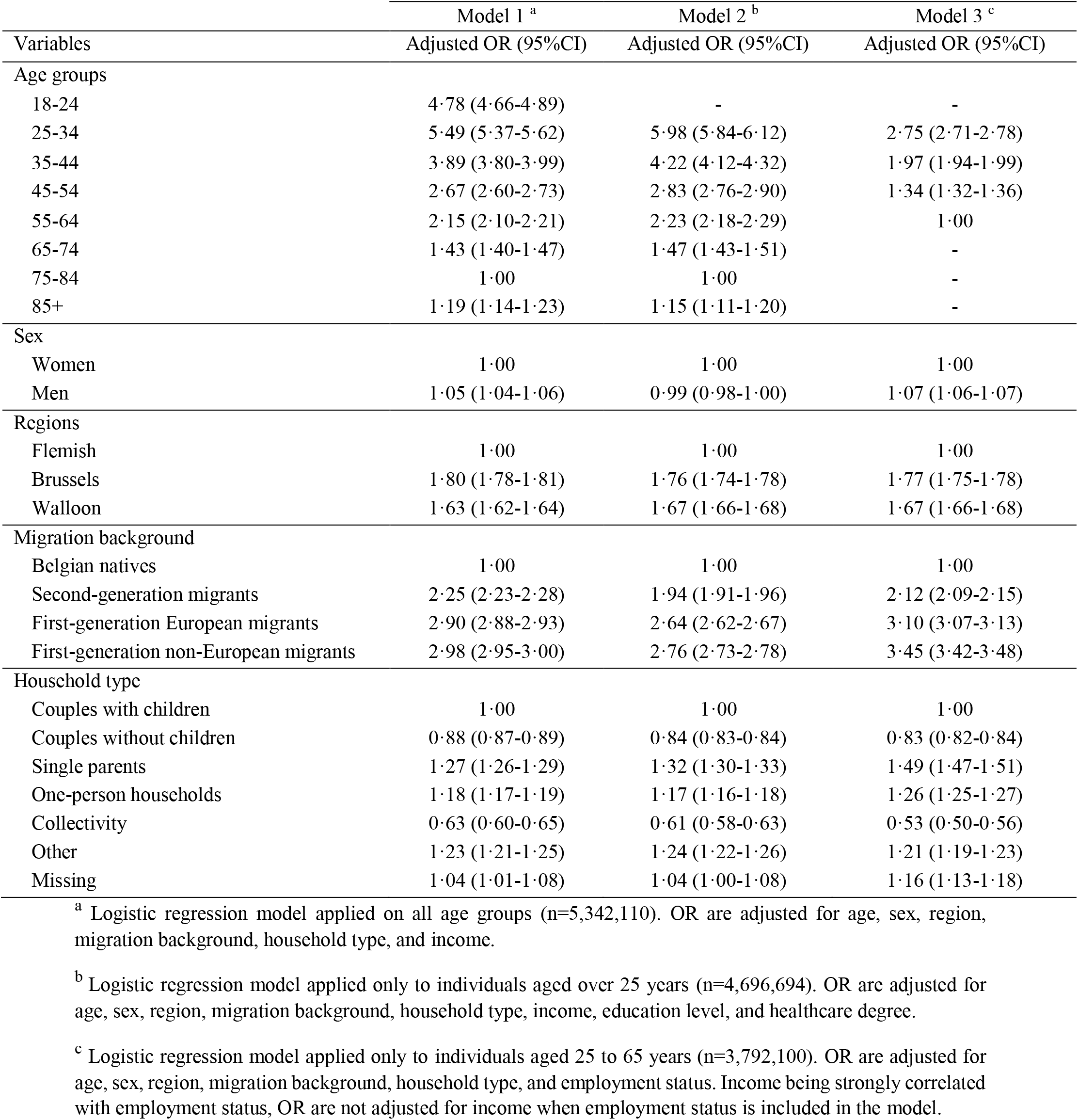
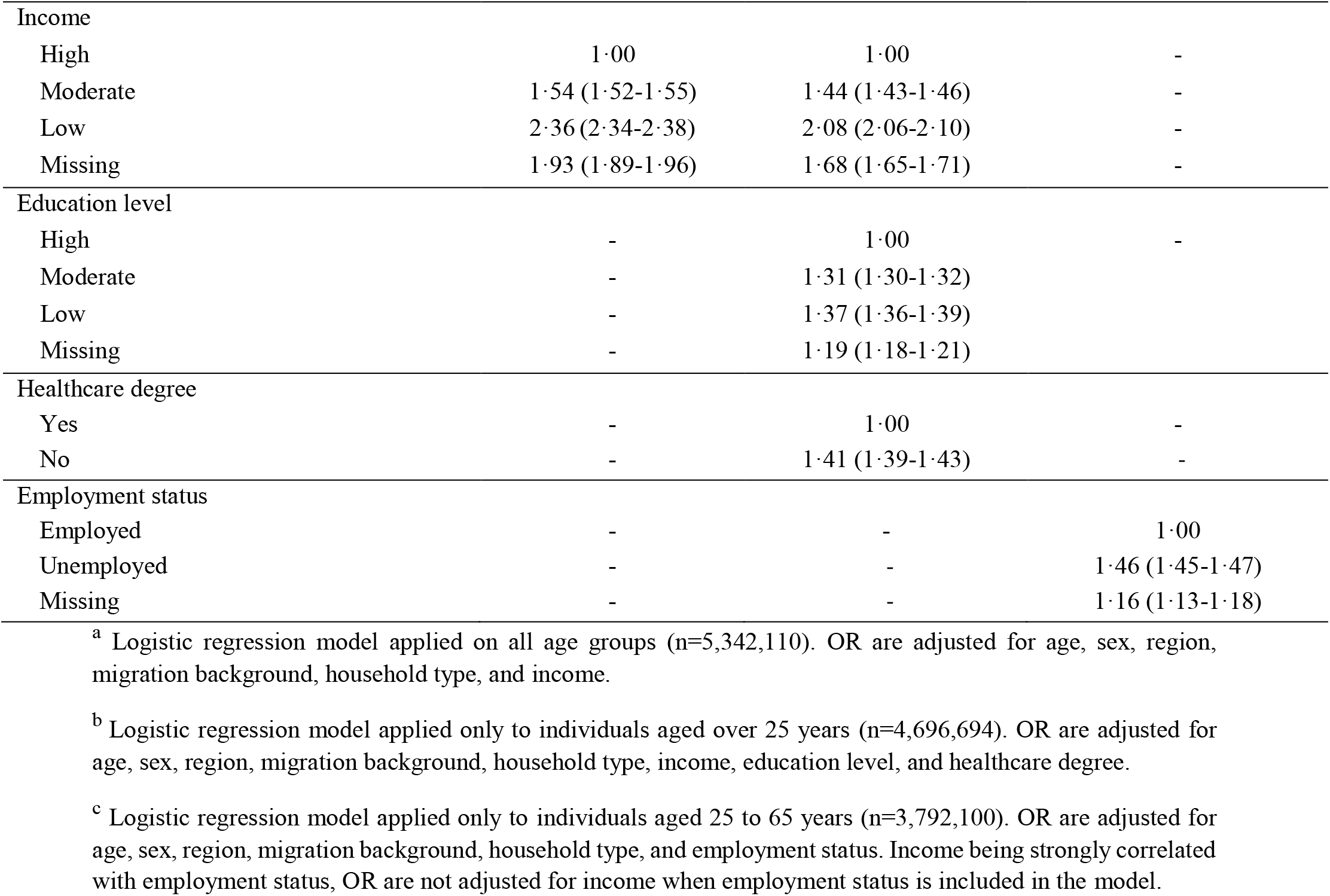
Adjusted odds ratio (OR) and their 95% confidence interval (CI) for the association between sociodemographic and socioeconomic characteristics and the odds of not having received a first dose of COVID-19 vaccine, Belgium, December 28^th^ 2020 - August 31^st^ 2021.

### Statistical analyses

The study population was first described with the number of individuals and corresponding percentages for all SD and SE characteristics according to the outcome (i.e. vaccination status for a first COVID-19 vaccine dose). To compute adjusted odds ratio (OR) and Wald 95% confidence intervals (95% CI), we fitted a first logistic regression model (Model 1) including age, sex, region, migration background, household type, and income as covariates. To assess the influence of education, a second model (Model 2) was fitted on a subset of our population comprising individuals aged 25-years and older adding education level and healthcare degree as additional covariates. Finally, to assess the impact of employment status, a third model (Model 3) was fitted on a subset of our population comprising individuals aged between 25- and 65-years adding employment status as a covariate and removing income (highly correlated variables).

Following the same procedures as those applied to the Belgian models, we fitted logistic regression models stratified by regions (Flanders, Brussels, Wallonia) based on the individual postal code of residence, in view of the important differences in vaccination coverage between them (see Table 1 and Figure 2). Furthermore, in Belgium, each regional health authority is responsible for the roll-out of the vaccination campaign and can therefore take its own measures for the practical implementation of the vaccination [14].

We performed sensitivity analyses to determine whether adding interaction terms between region and each covariate of the model, instead of stratifying by region, had an influence on the results. To do this, we extracted and compared, for both methods, the conditional probabilities of not having obtained a first COVID-19 vaccine dose according to all SD and SE characteristics (Supplementary Table 3).

All analyses were performed in R version 4.0.5 [19]. The package “Effects” was used to compute conditional probabilities [20].

## Results

### Study population

SD and SE characteristics of the study population are shown in Table 1. The final analyses included 5,342,110 individuals, representing 57·5% of the 2021 Belgian adult population (n=5,342,110/9,290,116). Among the study sample, there were only small proportional differences between men (46·7%) and women (53·4%). The largest age group consisted of those between 25 and 34 years (19·7%) and most individuals lived in the Flemish region (62·7%). In the 2021 Belgian adult population (see Supplementary Table 4), the proportion of women (51·2%) is also slightly higher than the proportion of men (48·8%). Its distribution of age groups differs slightly from that observed in our study sample. The largest age group in the Belgian adult population consist of those between 45 and 54 years (16·7%) and individuals aged between 25 and 34 years (16·1%) is the 4^th^ largest age group after individuals aged between 55 and 64 years (16·7%) and those aged between 35 and 44 years (16·2%). The Flemish region is also the most represented in the 2021 Belgian adult population although slightly over-represented in our study sample (58·2% versus 62·7%, respectively).

### Characteristics of unvaccinated individuals

Table 1 also shows the proportions of unvaccinated individuals in our entire sample according to all SD and SE characteristics. Among our study population, 536,716/5,342,110 (10·0%) Belgian residents did not receive a first dose of COVID-19 vaccine by August 31^st^ 2021. Overall, the proportion of unvaccinated individuals was inversely correlated with age, with 15.1% unvaccinated among 18–25-years-olds compared to 2·9% for those aged 85-years-olds and over. There was no notable difference in the proportion of unvaccinated individuals between women and men (10.0% and 10·1%, respectively). The proportion of unvaccinated individuals was higher in Brussels (22·0%), compared to Wallonia (12·0%) and Flanders 7·2%). Second-generation European migrants, first-generation European migrants, and first-generation non-European migrants had a higher proportion of unvaccinated individuals (19·1%, 18·9%, and 22·4%, respectively), compared to Belgian natives (5·2%). A higher proportion of unvaccinated individuals was identified among single parents (15·5%), couples with children (10·9%), and one-person households (10·5%), compared to collectivities (3·4%) and couples without children (5·6%). Proportionally more unvaccinated individuals were found in groups with a low education level (10·9%) and low income (15·5%), compared to individuals with a high education level (5·9%) and high income (5·1%). The proportion of unvaccinated between individuals employed (9·9%) and unemployed (9·6%) was essentially identical. The proportion of unvaccinated individuals was almost twice as high among those without a healthcare degree (10·4%), compared to those with a license (5·9%). Overall, it appeared that the unvaccinated individuals were over-represented in categories where data were indicated as missing (23·1% of unvaccinated for missing household type; 19·1% for missing education level; 17·3% for missing income; 24·4% for missing employment status).

### Predictors of first dose of COVID-19 vaccine uptake

Table 2 shows the adjusted OR estimated with logistic regression models to quantify the association between a lower uptake of a first dose of COVID-19 vaccine and SD/SE characteristics for Belgium. The multivariable results of the logistic regression models stratified by regions can be found in Supplementary Table 5. The crude OR for the Belgian models and those stratified by regions can be found in Supplementary Table 6 and Supplementary Table 7, respectively.

The multivariable results for Belgium (Table 2) show, in Model 1, including all age groups, an age gradient in COVID-19 vaccination coverage from 25-to 84-years-olds: the younger the individuals were, the more likely they were to be unvaccinated (OR 5·49 [5·37-5·62] for 25-34 age group, compared to 75-84 age group). Men had a slightly lower COVID-19 vaccine uptake, compared to women (OR 1·05 [1·04-1·06]). Individuals with a migration background had higher odds of being unvaccinated, compared to Belgian natives (OR 2·25 [2·23-2·28] for second-generation migrants; OR 2·90 [2·88-2·93] for first-generation European migrants; OR 2·98 [2·95-3·00] for first-generation non-European migrants). Compared to couples with children, one-person households (OR 1·18 [1·17-1·19) and single parents (OR 1·27 [1·26-1·29]) had higher odds of being unvaccinated. Individuals with missing information on the household type had also higher odds of being unvaccinated (OR 1·04 [1·01-1·08]). In terms of SE characteristics, a lower COVID-19 vaccine uptake was identified among individuals with low (OR 2·36 [2·34-2·38]), moderate (OR 1·54 [1·52-1·55]), or missing income (OR 1·93 [1·89-1·96]), compared to individuals with high income. In Model 2, including only individuals aged over 25-years-olds, a lower COVID-19 vaccine uptake was identified among individuals with a low (OR 1·37 [1·36-1·39]), moderate (OR 1·31 [1·30-1·32]), or missing (OR 1·19 [1·18-1·21]) education level, compared to individuals with a high education level. Not having a health care degree was also associated with a lower COVID-19 vaccine uptake (OR 1·41 [1·39-1·43]). In Model 3, including only individuals aged between 25- and 65 years-olds, being unemployed was associated with a higher odds of being unvaccinated (OR 1·46 [1·45-1·47]) as well as having a missing employment status (OR 1·16 [1·13-1·18]).

Overall, similar patterns could be identified in all regions with some exceptions regarding the Model 1 (Supplementary Table 5). In Flanders, the first-generation European migrants were the most likely to be unvaccinated with an OR of 4·35 [4·29-4·40] whereas considering Belgium overall, the first-generation non-European migrants had the largest OR (2·98 [2·95-3·00]). While in Belgium, one-person households were more likely to be unvaccinated compared to couples with children (OR 1·18 [1·17-1·19), in Brussels they were slightly less likely to be unvaccinated (OR 0·90 [0·89-0·92]). Moreover, in Belgium, the 25-34 age group had the highest odds of being unvaccinated (OR 5·49 [5·37-5·62]), whereas in Brussels, it was the 18-24 age group (OR 5·01 [4·73-5·32]).

## Discussion

Although only 34% of the Belgian population stated that they were definitively ready to be vaccinated against COVID-19 before the start of the vaccination campaign (study conducted between October 6 and 16, 2020) [21], Belgium had the 7^th^ highest coverage of the primary vaccination course in the European Union (89% of individuals over 18 years old had completed their primary course on 8 April 2022) [14,15]. Nevertheless, despite this high rate and free vaccination, we identified important SD and SE disparities in the uptake of a first COVID-19 vaccine dose in the Belgian adult population.

Overall, 10% of the individuals included in our study population did not receive a first dose of COVID-19 vaccine. A lower COVID-19 vaccine uptake was found among young individuals, men, migrants, single parents, one-person households, disadvantaged socioeconomic groups (with lower levels of income and education, unemployed), and individuals without a healthcare degree. A lower COVID-19 vaccine uptake was also observed among individuals for whom some SD/SE related variables (i.e., household type, income, education level, and employment status) were missing in the national registers. Overall, similar patterns could be identified in each region.

Our results are in line with those obtained in some other studies, conducted in different European and non-European countries, assessing SD and SE determinants associated with COVID-19 vaccine uptake in the general adult population. In Sweden, Spetz et al. found that COVID-19 vaccination coverage was lower among younger age groups and individuals having low income, living alone, and being born outside Sweden [9]. They found a gap in the COVID-19 vaccination coverage between men and women (with a 16% higher vaccination coverage among women), whereas in our study, men were only slightly less vaccinated than women. A cross-sectional study conducted in Wales identified that men, living in more deprived areas or urban areas, and ethnic groups other than white had higher odds of being unvaccinated [22]. In the United States, Williams et al. found that all racial and ethnic minority groups (except for Asians), compared with white group, had a lower COVID-19 vaccine uptake. They highlighted that age and SE factors (such as health insurance, income, education, employment) accounted for a large proportion of these social disparities and were therefore important key factors [10]. Our findings also partially confirm those of Barry et al. who identified, via aggregated national data, that COVID-19 vaccination coverage was lower among single parents, individuals with disabilities, and those living in counties with lower SE status and with a high percentage of households with children [23]. Our results, showing a higher vaccination coverage among individuals with a healthcare degree, are in line with Farah et al. who found a higher initial vaccination coverage among healthcare workers compared to the general population national average in the United States [24]. A cohort study conducted by Azamgarhi et al. during a time of high community COVID-19 prevalence in the United Kingdom also reported high early vaccination rates among healthcare workers [25].

Due to the rapid development of the COVID-19 vaccine, its efficacy and safety may have been undermined, leading to a higher level of mistrust of its benefits and concerns about its side effects, which are the strongest predictors of COVID-19 vaccine uptake [13,26,27]. Prior to the implementation of COVID-19 vaccination, numerous studies have investigated the SD and SE determinants associated with vaccine hesitancy that may lead to a lower intention to get the vaccine. Some have shown that being a female and having children were associated with a lower acceptance of the COVID-19 vaccine [27–30]. This sex difference was not observed in our results which show a lower COVID-19 vaccine uptake among men, compared to women. However, we similarly found that single parents had a lower COVID-19 vaccine uptake. A younger age was also identified as a predictor of lower intention to be vaccinated [27,28]. A higher vaccine hesitancy was identified among ethnic minority groups and SE disadvantaged groups (i.e. with lower levels of education and income, unemployment, and poor knowledge of COVID-19) [27–29]. This is again in line with our results showing a lower COVID-19 vaccine uptake among non-European migrants and SE disadvantaged groups. Vaccine hesitancy in migrants and SE disadvantaged groups may be underlined by several factors: in view of the more severe direct and indirect impact that the crisis has had on them (e.g. higher rate of COVID-19 infection, subsequent negative health outcomes, and unemployment) this may have increased distrust of governments, healthcare systems, and immunization [2,3,31,32]; decreased willingness to participate in public health measures as a result of decreased access to healthcare and resources [32]; raised concerns and negative assumptions about vaccination due to a lack of health literacy and recognition of misinformation that may be accentuated by the introduction of a vaccine based on novel technology [32–34]. Finally, although there was not a complete absence of fear of vaccination among healthcare workers, Wang et al. shown that they were less hesitant to be vaccinated, compared to non-healthcare workers [35]. In Belgium, before the start of the vaccination campaign, Kessel et al. [21] studied, through a survey conducted on a sample of 2,060 adults representative of the Belgian population, the predictors of willingness to be vaccinated against COVID-19. During the period in which the study was conducted (October 6 to 16, 2020), they did not identify social disparities in vaccine hesitancy by finding no association with education level, financial difficulties, and unemployment. In addition, they identified that men, compared to women, were more willing to get the COVID-19 vaccine. They also found that young individuals were more hesitant to be vaccinated.

It is important to note that other factors than hesitancy, due to fear or mistrust of COVID-19 vaccines, may explain the social disparities in vaccine uptake identified in our study. For example, administrative hurdles may partly explain the lower coverage in some SE groups. Indeed, our results showed a lower vaccine uptake among individuals whose SE information was partly missing from national registers. For most individuals in Belgium, the vaccination process was initiated after receiving an invitation sent by the regions and being absent of some registers may be indicative of a hard-to-reach population in the context of a broad automated invitation process. Language barriers may also be another factor explaining the lower COVID-19 vaccination coverage among migrants who may have difficulty with the vaccination related information primarily available in the Belgian official languages.

To our knowledge, this paper is the first large representative study investigating, thanks to an individual data linkage established within the LINK-VACC project, SD and SE disparities in vaccination coverage in Belgium and all regions with up-to-date SD and SE information. Our study has several limitations. First, individuals never tested for COVID-19 before August 31^st^ 2021 are not included in our study population. Indeed, because of the General Data Protection Regulation, individual’s SD/SE information from the DEMOBEL database could not be obtained from the total Belgian population, the master database used for the linkage consists of the COVID-19 Healthdata test database obtained on the 31^st^ of August 2021. This resulted in a slight over-representation of vaccinated individuals in our study population compared to the actual vaccination coverage in Belgium (90·0% vs 85.4%, respectively). In view of this gap, one could assume that untested individuals who are more likely to be unvaccinated also belong to more SE disadvantaged groups. This would make our results more conservative because inequalities in vaccination are certainly wider than those observed in our study. However, despite this slight over-representation of vaccinated individuals in our study population, it should be pointed out that the trends in vaccination coverage per region, age group, and sex are very similar to those of the overall Belgian adult population (see Supplementary Figure 1). A second limitation is that our study sample does not include the unregistered population, a group of about 100,000-150,000 individuals. Third, individuals vaccinated abroad are not automatically registered in Vaccinnet+ (e.g. frontier workers). This could partly explain the lower COVID-19 vaccine uptake among the first-generation European migrants in Flanders. Fourth, we only looked at inequity in COVID-19 vaccine uptake and we cannot identify whether this results from personal conviction or inequity in the distribution of the vaccine, which is a necessary step to investigate to improve the vaccination campaigns.

Vaccination being still on-going at the time of the study, the definition of vaccinated individuals aimed at capturing those who were readily reached by the vaccination campaign and decided reasonably quickly to get vaccinated (e.g. as shown in Figure 1, vaccination coverage of 18-24-years-olds after August 31^st^ 2021 is still increasing very slowly). Future research could also include individuals who were vaccinated later, vaccination among adolescents and children, or factors influencing the uptake of the booster. Another perspective would be to investigate the factors underlying the refusal to be vaccinated in Belgium, specifically among the groups highlighted by our study as having a lower COVID-19 vaccine uptake and identified by other studies as being more likely to be infected by the SARS-CoV-2 or to develop severe forms of the disease (e.g. SE disadvantaged groups).

## Conclusions

Despite the success of the vaccination campaign in Belgium (89% of adults vaccinated with primary course), free vaccination, and the efforts made by the regional health authorities to reach all citizens, important SD and SE inequalities in COVID-19 vaccine uptake were identified. Our study contributes to a better identification of disparities in COVID-19 vaccination and helps to better target vaccination strategies to more vulnerable groups with the goal of acquiring the broadest possible vaccine coverage to limit the circulation of the virus and the development of severe negative health outcomes, whether in the context of the COVID-19 pandemic or other potential health threats.

## Data Availability

The data used in this study are pseudonymised individual data obtained from several national registers hosted by Healthdata.be, an independent institution within Sciensano aiming to bring together all the data currently stored in multiple health registers on a single web-based platform. Due to the General Data Protection Regulation (GDPR) legislations in Belgium, these data are not publicly available. Data request must be addressed to the Information Security Committee (ISC).

## List of abbreviations

SARS-CoV-2: Severe Acute Respiratory Syndrome Coronavirus 2
COVID-19: Coronavirus disease 2019
SD: Sociodemographic
SE: Socioeconomic
Statbel: Statistics Belgium
CoBRHA: Common Base Registry for HealthCare Actors
ISCED: International Standard Classification of Education.

## Declarations

### Ethics approval and consent to participate

The protocol of the LINK-VACC project was approved by the Medical Ethics Committee from the Vrije Universiteit of Brussels on 3 February 2021 (B.U.N 1432020000371) and obtained authorization from the Information Security Committee (ISC) Social Security and Health (reference number: IVC/KSZG/21/034). The study protocol has been preregistered on ClinicalTrials.gov (ClinicalTrials.gov ID: NCT05373420).

As confirmed by sections 23 and 24 of the *Guidelines 03/2020 on the processing of data concerning health for the purpose of scientific research in the context of the COVID-19 outbreak* of the European Data Protection Board (V1.0 of 21 April 2020), this survey falls under Article 6 §1(e) and Article 9 §2(i) of the General Data Protection Regulation (GDPR). In appliance with these GDPR legal grounds of data processing, no informed consent had to be signed by the patients.

### Consent for publication

Not applicable

### Availability of data and materials

The data used in this study are pseudonymised individual data obtained from several national registers hosted by Healthdata.be, an independent institution within Sciensano aiming to bring together all the data currently stored in multiple health registers on a single web-based platform. Due to the General Data Protection Regulation (GDPR) legislations in Belgium, these data are not publicly available. Data request must be addressed to the Information Security Committee (ISC). The code used in these analyses can be provided upon request.

## Authors’ contributions

LiC reviewed the literature; LiC, JvL, BDV, LVdB, NS, RDB, LuC, and PH conceived the study; LiC, JvL, LVdB, RDB, LuC, and PH selected the population; LiC and PH reviewed all available data; LiC, JvL, BDV, LVdB, NS, RDB, and PH designed the statistical methodology; LiC conducted the statistical analyses; LiC, JvL, LVdB, RDB, and PH interpreted the findings; LiC and PH designed the figures; LiC wrote the first draft of the paper. All authors revised the text. LiC, JvL, MB, VS, LuC, and PH have directly access to the data of the present study. LiC, JvL, MB, VS, LuC, and PH have full access to all the data in the study. LiC and PH has verified the underlying data of the study. All authors approved the final version of this manuscript and accepted responsibility for its submission for publication.

## Acknowledgments

We are very grateful to Statistics Belgium for their essential advice on the recent socioeconomic data. We would also like to thank Healthdata and the Belgian regions.

