## Additional file 1 for "Sociodemographic and socioeconomic disparities in COVID-19 vaccine uptake in Belgium – A nationwide record linkage study"

* Correspondence **Lisa Cavillot**

Sciensano

Rue Ernest Blerot 1

1070 Anderlecht

Belgique

Tel : +32475617195

### Supplementary Figures

#### Supplementary Figure 1: Vaccination coverage of a first dose of COVID-19 vaccine according to region, age group, and sex in the 2021 Belgian adult population and in our study population including only adults tested at least once in Belgium by August 31^st^ 2021, Belgium, December 28^th^ 2020 - August 31^st^ 2021.

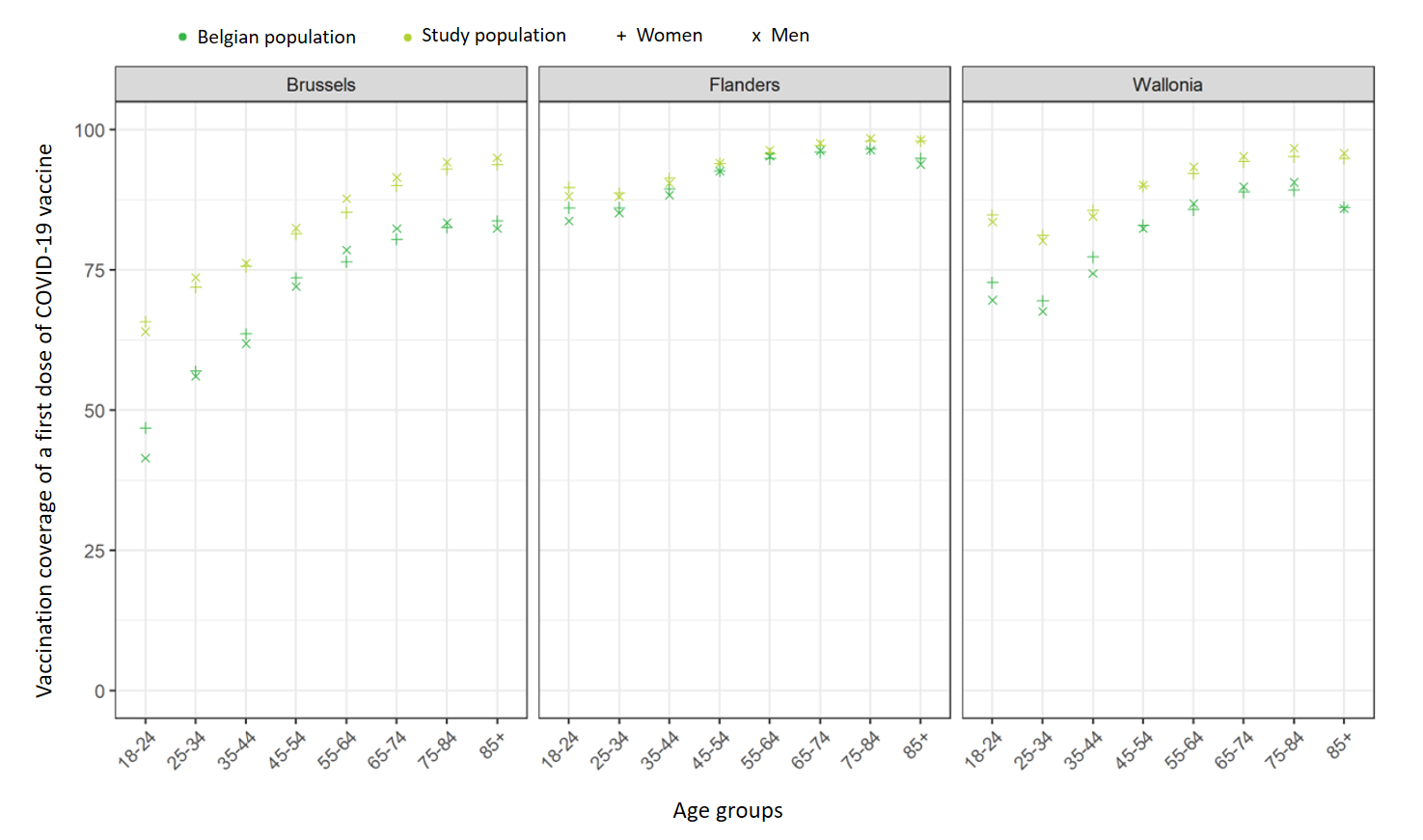

### Supplementary Tables

#### Supplementary Table 1: Data holder and time frame for all sociodemographic and socioeconomic variables included as covariates in the logistic regression models, Belgium, December 28^th^ 2020 - August 31^st^ 2021.

| **Variables** | **Data holder** | **Time frame** |
| --- | --- | --- |
| Age | National register | March 2022 |
| Sex | National register | March 2022 |
| Region | National register | March 2022 |
| Migration background | Statistics Belgium | August 2021 |
| Household type | Statistics Belgium | August 2021 |
| Income | Statistics Belgium | 2018 (fiscal year 2019) |
| Education level | Statistics Belgium | Census 2017 |
| Employment status | Statistics Belgium | Year 2019 |
| Healthcare degree | Common Base Registry for HealthCare Actor | January 2021 |

#### Supplementary Table 2: Characterization of the population with at least one missing information regarding household type, education level, income, or employment status relative to the total population by each sociodemographic and socioeconomic characteristic.

| **Variables** | **Total**  **(n=5,342,110)** | **At least one missing information (n=764,857)** | **At least one missing information to the total population** |
| --- | --- | --- | --- |
|  | n | n | %_row_ |
| Age groups (years) |  |  |  |
| 18-24 | 645,416 | 55,991 | 8·66 |
| 25-34 | 1,051,576 | 182,302 | 17·34 |
| 35-44 | 978,478 | 192,936 | 19·72 |
| 45-54 | 903,514 | 133,289 | 14·75 |
| 55-64 | 797,786 | 76,903 | 9·64 |
| 65-74 | 511,784 | 46,103 | 9·01 |
| 75-84 | 296,974 | 37,353 | 12·58 |
| 85+ | 156,582 | 39,980 | 25·53 |
| Regions |  |  |  |
| Brussels | 579,687 | 205,993 | 35·53 |
| Flemish | 3,348,547 | 392,471 | 11·72 |
| Walloon | 1,413,876 | 166,393 | 11·77 |
| Sex |  |  |  |
| Women | 2,849,932 | 401,457 | 14·09 |
| Men | 2,492,178 | 363,400 | 14·58 |
| Migration background |  |  |  |
| Belgian natives | 3,652,357 | 181,359 | 4·97 |
| Second-generation migrants | 324,420 | 13,868 | 4·27 |
| First-generation European migrants | 607,586 | 265,626 | 43·72 |
| First-generation non-European migrants | 757,747 | 304,004 | 40·12 |
| Household type |  |  |  |
| One person households | 878,506 | 135,337 | 15·41 |
| Collectivity | 96,213 | 67,015 | 69·65 |
| Couples without children | 1,302,576 | 124,823 | 9·58 |
| Couples with children | 2,363,467 | 315,030 | 13·33 |
| Single parents | 548,369 | 61,662 | 11·24 |
| Missing | 26,752 | 26,752 | 100 |
| Other | 126,227 | 34,238 | 27·12 |
| Education level |  |  |  |
| Low | 1,567,744 | 44,076 | 2·81 |
| Moderate | 1,569,361 | 15,132 | 0·96 |
| High | 1,511,494 | 12,138 | 0·80 |
| Missing | 693,511 | 693,511 | 100 |
| Income |  |  |  |
| Low | 1,815,339 | 352,284 | 19·41 |
| Moderate | 1,564,464 | 126,673 | 8·10 |
| High | 1,762,607 | 86,200 | 4·89 |
| Missing | 199,700 | 199,700 | 100 |
| Healthcare degree |  |  |  |
| No | 4,918,258 | 738,114 | 15·01 |
| Yes | 423,852 | 26,743 | 6·31 |
| Employment status |  |  |  |
| Unemployed | 1,874,544 | 277,670 | 14·81 |
| Employed | 3,364,795 | 384,416 | 11·42 |
| Missing | 102,771 | 102,771 | 100 |

n: number of individuals; %row: percentage in row

#### Supplementary Table 3: Conditional probabilities of not having receive a first dose of COVID-19 vaccine obtained by logistic regression models stratified by region and logistic regression models including region as an interaction term between each sociodemographic and socioeconomic characteristic, Belgium, December 28^th^ 2020 - August 31^st^ 2021.

|  | **Flanders** | | **Brussels** | | **Wallonia** | |
| --- | --- | --- | --- | --- | --- | --- |
| **Variables** | **Probabilities (IC95%)** | | **Probabilities (IC95%)** | | **Probabilities (IC95%)** | |
|  | **Stratification** | **Interaction** | **Stratification** | **Interaction** | **Stratification** | **Interaction** |
| Age (years) ^a^ |  |  |  |  |  |  |
| 18-24 | 0·076 (0·075-0·077) | 0·086 (0·085-0·087) | 0·301 (0·298-0·305) | 0·197 (0·194-0·201) | 0·133 (0·132-0·135) | 0·133 (0·131-0·135) |
| 25-34 | 0·089 (0·088-0·090) | 0·010 (0·099-0·101) | 0·262 (0·260-0·264) | 0·169 (0·167-0·171) | 0·176 (0·175-0·178) | 0·176 (0·175-0·178) |
| 35-44 | 0·065 (0·064-0·065) | 0·073 (0·072-0·074) | 0·203 (0·201-0·206) | 0·129 (0·127-0·131) | 0·129 (0·128-0·131) | 0·129 (0·128-0·130) |
| 45-54 | 0·046 (0·045-0·047) | 0·052 (0·051-0·052) | 0·150 (0·148-0·152) | 0·093 (0·091-0·095) | 0·089 (0·088-0·090) | 0·089 (0·087-0·090) |
| 55-64 | 0·037 (0·036-0·038) | 0·041 (0·041-0·042) | 0·130 (0·127-0·132) | 0·079 (0·077-0·081) | 0·071 (0·070-0·072) | 0·071 (0·070-0·072) |
| 65-74 | 0·023 (0·022-0·023) | 0·026 (0·025-0·027) | 0·101 (0·098-0·104) | 0·061 (0·059-0·062) | 0·049 (0·048-0·051) | 0·050 (0·048-0·051) |
| 75-84 | 0·015 (0·014-0·015) | 0·017 (0·016-0·018) | 0·078 (0·075-0·083) | 0·047 (0·044-0·049) | 0·038 (0·036-0·039) | 0·038 (0·036-0·039) |
| 85+ | 0·017 (0·016-0·018) | 0·020 (0·019-0·020) | 0·087 (0·081-0·093) | 0·052 (0·048-0·056) | 0·046 (0·044-0·049) | 0·046 (0·044-0·049) |
| Sex ^a^ |  |  |  |  |  |  |
| Women | 0·047 (0·046-0·047) | 0·053 (0·054-0·057) | 0·190 (0·188-0·191) | 0·110 (0·108-0·111) | 0·096 (0·096-0·097) | 0·097 (0·096-0·102) |
| Men | 0·050 (0·049-0·050) | 0·057 (0·057-0·059) | 0·188 (0·186-0·189) | 0·109 (0·107-0·110) | 0·101 (0·100-0·102) | 0·101 (0·101-0·102) |
| Migration background ^a^ |  |  |  |  |  |  |
| Belgian natives | 0·036 (0·036-0·037) | 0·038 (0·038-0·038) | 0·106 (0·104-0·107) | 0·084 (0·082-0·085) | 0·080 (0·079-0·080) | 0·080 (0·079-0·080) |
| Second generation migrants | 0·092 (0·090-0·093) | 0·096 (0·095-0·097) | 0·218 (0·215-0·222) | 0·178 (0·175-0·182) | 0·127 (0·126-0·129) | 0·128 (0·126-0·129) |
| First generation European migrants | 0·141 (0·140-0·143) | 0·147 (0·146-0·148) | 0·208 (0·206-0·210) | 0·169 (0·167-0·171) | 0·150 (0·149-0·152) | 0·150 (0·149-0·152) |
| First generation non-European migrants | 0·109 (0·107-0·110) | 0·114 (0·113-0·115) | 0·253 (0·251-0·255) | 0·208 (0·206-0·210) | 0·171 (0·169-0·173) | 0·171 (0·169-0·173) |

**Supplementary table 3: Continued**

|  | **Flanders** | | **Brussels** | | **Wallonia** | |
| --- | --- | --- | --- | --- | --- | --- |
| **Variables** | **Probabilities (IC95%)** | | **Probabilities (IC95%)** | | **Probabilities (IC95%)** | |
|  | **Stratification** | **Interaction** | **Stratification** | **Interaction** | **Stratification** | **Interaction** |
| Household type ^a^ |  |  |  |  |  |  |
| Couples with children | 0·046 (0·046-0·047) | 0·053 (0·052-0·053) | 0·202 (0·200-0·204) | 0·119 (0·118-0·121) | 0·094 (0·093-0·095) | 0·094 (0·094-0·095) |
| Couples | 0·043 (0·043-0·044) | 0·049 (0·049-0·050) | 0·153 (0·151-0·156) | 0·088 (0·087-0·090) | 0·089 (0·088-0·091) | 0·090 (0·089-0·091) |
| Single parents | 0·061 (0·061-0·062) | 0·070 (0·070-0·071) | 0·216 (0·213-0·219) | 0·129 (0·126-0·131) | 0·116 (0·115-0·118) | 0·117 (0·116-0·119) |
| One person households | 0·059 (0·058-0·060) | 0·067 (0·067-0·068) | 0·186 (0·184-0·188) | 0·109 (0·107-0·112) | 0·119 (0·117-0·120) | 0·119 (0·118-0·121) |
| Collectivity | 0·031 (0·030-0·033) | 0·035 (0·033-0·037) | 0·142 (0·131-0·153) | 0·083 (0·076-0·090) | 0·055 (0·052-0·058) | 0·055 (0·052-0·058) |
| Other | 0·064 (0·063-0·066) | 0·073 (0·071-0·074) | 0·178 (0·173-0·182) | 0·104 (0·102-0·108) | 0·125 (0·122-0·129) | 0·125 (0·121-0·129) |
| Missing | 0·046 (0·044-0·048) | 0·057 (0·054-0·059) | 0·181 (0·173-0·190) | 0·113 (0·107-0·119) | 0·076 (0·071-0·081) | 0·099 (0·093-0·105) |
| Income ^a^ |  |  |  |  |  |  |
| High | 0·032 (0·031-0·032) | 0·036 (0·036-0·036) | 0·109 (0·107-0·111) | 0·068 (0·067-0·070) | 0·068 (0·068-0·069) | 0·069 (0·069-0·070) |
| Moderate | 0·048 (0·048-0·049) | 0·054 (0·054-0·055) | 0·182 (0·180-0·185) | 0·119 (0·117-0·121) | 0·095 (0·094-0·096) | 0·096 (0·095-0·097) |
| Low | 0·075 (0·074-0·075) | 0·083 (0·083-0·084) | 0·234 (0·232-0·235) | 0·155 (0·153-0·157) | 0·137 (0·136-0·138) | 0·138 (0·137-0·139) |
| Missing | 0·068 (0·066-0·070) | 0·075 (0·073-0·076) | 0·183 (0·179-0·186) | 0·119 (0·116-0·122) | 0·122 (0·118-0·126) | 0·122 (0·119-0·126) |
| **Population aged 25 years and over (n=4,696,694)** | | | | | | |
| Education level ^b^ |  |  |  |  |  |  |
| High | 0·038 (0·037-0·038) | 0·043 (0·042-0·043) | 0·144 (0·141-0·146) | 0·082 (0·081-0·084) | 0·081 (0·080-0·082) | 0·081 (0·081-0·083) |
| Moderate | 0·049 (0·048-0·049) | 0·055 (0·055-0·056) | 0·213 (0·211-0·216) | 0·127 (0·124-0·129) | 0·096 (0·095-0·097) | 0·098 (0·096-0·098) |
| Low | 0·050 (0·049-0·051) | 0·057 (0·056-0·057) | 0·210 (0·208-0·213) | 0·125 (0·123-0·128) | 0·103 (0·102-0·104) | 0·104 (0·102-0·105) |
| Missing | 0·049 (0·048-0·050) | 0·055 (0·055-0·056) | 0·153 (0·151-0·155) | 0·088 (0·087-0·090) | 0·095 (0·094-0·097) | 0·096 (0·094-0·097) |
| Healthcare degree ^b^ |  |  |  |  |  |  |
| Yes | 0·034 (0·033-0·035) | 0·039 (0·038-0·039) | 0·124 (0·120-0·128) | 0·076 (0·073-0·078) | 0·071 (0·069-0·072) | 0·071 (0·069-0·072) |
| No | 0·046 (0·046-0·046) | 0·053 (0·052-0·053) | 0·174 (0·172-0·175) | 0·108 (0·107-0·109) | 0·095 (0·095-0·096) | 0·095 (0·095-0·096) |

**Supplementary table 3: Continued**

|  | **Flanders** | | **Brussels** | | **Wallonia** | |
| --- | --- | --- | --- | --- | --- | --- |
| **Variables** | **Probabilities (IC95%)** | | **Probabilities (IC95%)** | | **Probabilities (IC95%)** | |
|  | **Stratification** | **Stratification** | **Stratification** | **Interaction** | **Stratification** | **Interaction** |
| **Population aged between 25 and 65 years (n=3,792,100)** | | | | | | |
| Employment status ^c^ |  |  |  |  |  |  |
| Employed | 0·055 (0·055-0·056) | 0·062 (0·062-0·063) | 0·193 (0·191-0·194) | 0·122 (0·120-0·123) | 0·109 (0·108-0·109) | 0·111 (0·110-0·111) |
| Unemployed | 0·084 (0·083-0·085) | 0·094 (0·093-0·095) | 0·242 (0·239-0·244) | 0·155 (0·153-0·158) | 0·146 (0·144-0·147) | 0·149 (0·147-0·150) |
| Missing | 0·074 (0·072-0·077) | 0·082 (0·080-0·084) | 0·184 (0·179-0·189) | 0·116 (0·112-0·120) | 0·134 (0·129-0·140) | 0·136 (0·130-0·141) |

^a^ Logistic regression model applied on all age groups (Flanders, n=3,348,547; Brussel, n=579,687; Wallonia, n=1,413,876). Probabilities are adjusted for age, sex, region, migration background, household type, and income.

^b^ Logistic regression model applied only to individuals aged over 25 years (Flanders, n=2,955,807; Brussel, n=505,082; Wallonia, n=1,235,805). Probabilities are adjusted for age, sex, region, migration background, household type, income, education level, and healthcare degree.

^c^ Logistic regression model applied only to individuals aged 25 to 65 years (Flanders, n=2,357,375; Brussel, n=436,356; Wallonia, n=998,369). Probabilities are adjusted for age, sex, region, migration background, household type, and employment status. Income being strongly correlated with employment status, probabilities are not adjusted for income when employment status is included in the model.

#### Supplementary Table 4: Distribution of age, region, and sex in the 2021 Belgian adult population according to the uptake of a first dose of COVID-19 vaccine, Belgium, 2021.

| **Variables** | **All (n=9,209,116)** | **First dose not administered (n=1,345,623)** | **First dose administered (n=7,863,493)** |
| --- | --- | --- | --- |
|  | **n (%_column_)** | **n (%_row_)** | **n (%_row_)** |
| Age groups (years) |  |  |  |
| 18-24 | 922,803 (10·02) | 224,320 (24·31) | 698,482 (75·69) |
| 25-34 | 1,486,440 (16·14) | 352,963 (23·75) | 1,133,477 (76·25) |
| 35-44 | 1,493,556 (16·22) | 274,471 (18·38) | 1,219,085 (81·62) |
| 45-54 | 1,538,889 (16·71) | 194,227 (12·62) | 1,344,662 (87·38) |
| 55-64 | 1,538,307 (16·70) | 140,347 (9·12) | 1,397,960 (90·88) |
| 65-74 | 1,194,091 (12·97) | 85,086 (7·13) | 1,109,005 (92·87) |
| 75-84 | 703,107 (7·63) | 45,018 (6·40) | 658,089 (93·60) |
| 85+ | 331,923 (3·60) | 29,190 (8·79) | 302,733 (91·21) |
| Regions |  |  |  |
| Brussels | 944,417 (10·25) | 318,944 (33·77) | 625,473 (66·23) |
| Flemish | 5,363,075 (58·24) | 454,399 (8·47) | 4,908,676 (91·53) |
| Walloon | 2,901,624 (31·51) | 572,280 (19·72) | 2,329,344 (80·28) |
| Sex |  |  |  |
| Women | 4,714,364 (51·19) | 667,521 (14·16) | 4,046 ,843 (85·84) |
| Men | 4,494,752 (48·81) | 678,102 (15·09) | 3,816,650 (84·91) |

#### Supplementary Table 5: Adjusted OR and their 95% confidence interval (CI) for the association between sociodemographic and socioeconomic characteristics and the odds of not having received a first dose of COVID-19 vaccine stratified by region, Belgium, December 28^th^ 2020 - August 31^st^ 2021.

|  | **Flanders** | | | **Brussels** | | | **Wallonia** | | |
| --- | --- | --- | --- | --- | --- | --- | --- | --- | --- |
| **Variables** | **Adjusted OR (95%CI)** | | | **Adjusted OR (95% CI)** | | | **Adjusted OR (95%CI)** | | |
|  | **Model 1 ^a^** | **Model 2 ^b^** | **Model 3 ^c^** | **Model 1 ^a^** | **Model 2 ^b^** | **Model 3 ^c^** | **Model 1 ^a^** | **Model 2 ^b^** | **Model 3 ^c^** |
| Age (years) |  |  |  |  |  |  |  |  |  |
| 18-24 | 5·46 (5·26-5·66) | - | - | 5·01 (4·73-5·32) | - | - | 3·91 (3·75-4·07) | - | - |
| 25-34 | 6·46 (6·24-6·69) | 7·02 (6·77-7·27) | 2·79 (2·74-2·83) | 4·15 (3·92-4·39) | 4·64 (4·38-4·91) | 2·44 (2·38-2·50) | 5·45 (5·24-5·67) | 5·70 (5·48-5·93) | 3·03 (2·97-3·09) |
| 35-44 | 4·57 (4·41-4·74) | 4·91 (4·73-5·09) | 2·02 (1·98-2·05) | 3·01 (2·85-3·19) | 3·48 (3·39-3·69) | 1·76 (1·72-1·81) | 3·77 (3·63-3·93) | 3·97 (3·81-4·13) | 2·11 (2·06-2·15) |
| 45-54 | 3·19 (3·07-3·30) | 3·35 (3·23-3·47) | 1·39 (1·36-1·41) | 2·08 (1·97-2·21) | 2·34 (2·21-2·48) | 1·24 (1·20-1·27) | 2·47 (2·38-2·57) | 2·58 (2·47-2·68) | 1·37 (1·34-1·40) |
| 55-64 | 2·51 (2·43-2·61) | 2·60 (2·51-2·70) | 1·00 | 1·75 (1·65-1·86) | 1·88 (1·77-1·99) | 1·00 | 1·94 (1·87-2·02) | 1·99 (1·91-2·08) | 1·00 |
| 65-74 | 1·55 (1·49-1·61) | 1·59 (1·53-1·66) | - | 1·32 (1·24-1·41) | 1·36 (1·28-1·45) | - | 1·32 (1·27-1·38) | 1·35 (1·29-1·41) | - |
| 75-84 | 1·00 | 1·00 | - | 1·00 | 1·00 | - | 1·00 | 1·00 | - |
| 85+ | 1·16 (1·10-1·23) | 1·13 (1·07-1·19) | - | 1·12 (1·02-1·24) | 1·07 (0·97-1·17) | - | 1·24 (1·17-1·32) | 1·21 (1·14-1·29) | - |
| Sex |  |  |  |  |  |  |  |  |  |
| Women | 1·00 | 1·00 | 1·00 | 1·00 | 1·00 | 1·00 | 1·00 | 1·00 | 1·00 |
| Men | 1·07 (1·06-1·08) | 1·00 (0·99-1·01) | 1·08 (1·07-1·10) | 0·99 (0·98-1·00) | 0·94 (0·93-0·96) | 1·01 (1·00-1·03) | 1·05 (1·04-1·06) | 0·99 (0·98-1·01) | 1·06 (1·05-1·08) |
| Migration background |  |  |  |  |  |  |  |  |  |
| Belgian natives | 1·00 | 1·00 | 1·00 | 1·00 | 1·00 | 1·00 | 1·00 | 1·00 | 1·00 |
| Second generation migrants | 2·68 (2·63-2·72) | 2·33 (2·28-2·38) | 2·55 (2·49-2·60) | 2·37 (2·31-2·44) | 1·83 (1·77-1·90) | 2·20 (2·12-2·28) | 1·69 (1·66-1·72) | 1·54 (1·51-1·57) | 1·62 (1·59-1·66) |
| First-generation European migrants | 4·35 (4·29-4·40) | 3·77 (3·71-3·82) | 4·76 (4·70-4·83) | 2·22 (2·17-2·27) | 2·14 (2·08-2·19) | 2·45 (2·39-2·51) | 2·04 (2·01-2·07) | 1·90 (1·87-1·93) | 2·11 (2·07-2·14) |
| First-generation non-European migrants | 3·23 (3·20-3·27) | 2·83 (2·79-2·87) | 3·75 (3·70-3·79) | 2·87 (2·81-2·93) | 2·63 (2·57-2·70) | 3·51 (3·43-3·60) | 2·38 (2·34-2·41) | 2·24 (2·21-2·28) | 2·62 (2·58-2·66) |

**Supplementary Table 5: Continued**

|  | **Flanders** | | | **Brussels** | | | **Wallonia** | | |
| --- | --- | --- | --- | --- | --- | --- | --- | --- | --- |
| **Variables** | **Adjusted OR (95%CI)** | | | **Adjusted OR (95% CI)** | | | **Adjusted OR (95%CI)** | | |
|  | **Model 1 ^a^** | **Model 2 ^b^** | **Model 3 ^c^** | **Model 1 ^a^** | **Model 2 ^b^** | **Model 3 ^c^** | **Model 1 ^a^** | **Model 2 ^b^** | **Model 3 ^c^** |
| Household type |  |  |  |  |  |  |  |  |  |
| Couples with children | 1·00 | 1·00 | 1·00 | 1·00 | 1·00 | 1·00 | 1·00 | 1·00 | 1·00 |
| Couples without children | 0·93 (0·92-0·94) | 0·87 (0·86-0·89) | 0·87 (0·86-0·88) | 0·72 (0·70-0·73) | 0·71 (0·69-0·73) | 0·67 (0·65-0·69) | 0·95 (0·93-0·96) | 0·89 (0·88-0·91) | 0·90 (0·88-0·92) |
| Single parents | 1·35 (1·33-1·37) | 1·41 (1·39-1·44) | 1·60 (1·57-1·62) | 1·09 (1·07-1·11) | 1·13 (1·10-1·15) | 1·25 (1·22-1·28) | 1·27 (1·25-1·29) | 1·30 (1·27-1·32) | 1·47 (1·44-1·49) |
| One-person households | 1·29 (1·27-1·31) | 1·27 (1·25-1·29) | 1·39 (1·37-1·41) | 0·90 (0·89-0·92) | 0·92 (0·90-0·93) | 0·93 (0·92-0·95) | 1·30 (1·28-1·32) | 1·26 (1·24-1·28) | 1·39 (1·37-1·42) |
| Collectivity | 0·65 (0·62-0·69) | 0·64 (0·60-0·67) | 0·58 (0·54-0·63) | 0·67 (0·61-0·73) | 0·63 (0·57-0·69) | 0·56 (0·49-0·63) | 0·56 (0·52-0·60) | 0·55 (0·51-0·59) | 0·46 (0·42-0·51) |
| Other | 1·41 (1·37-1·44) | 1·42 (1·38-1·46) | 1·41 (1·37-1·45) | 0·86 (0·84-0·89) | 0·89 (0·86-0·92) | 0·83 (0·80-0·86) | 1·37 (1·32-1·42) | 1·33 (1·28-1·38) | 1·30 (1·25-1·36) |
| Missing | 1·08 (1·03-1·13) | 1·09 (1·03-1·15) | 1·03 (0·97-1·09) | 0·94 (0·89-0·99) | 0·93 (0·87-0·99) | 0·94 (0·91-0·98) | 1·06 (0·99-1·13) | 1·07 (0·99-1·16) | 1·00 (0·92-1·09) |
| Income |  |  |  |  |  |  |  |  |  |
| High | 1·00 | 1·00 | - | 1·00 | 1·00 | - | 1·00 | 1·00 | - |
| Moderate | 1·54 (1·52-1·56) | 1·45 (1·43-1·47) | - | 1·84 (1·79-1·88) | 1·66 (1·62-1·71) | - | 1·43 (1·41-1·45) | 1·35 (1·32-1·37) | - |
| Low | 2·44 (2·41-2·47) | 2·17 (2·14-2·20) | - | 2·51 (2·45-2·57) | 2·17 (2·12-2·23) | - | 2·15 (2·12-2·18) | 1·91 (1·88-1·94) | - |
| Missing | 2·17 (2·12-2·22) | 1·84 (1·79-1·90) | - | 1·83 (1·78-1·89) | 1·69 (1·63-1·75) | - | 1·87 (1·81-1·94) | 1·61 (1·55-1·68) | - |
| Education level |  |  |  |  |  |  |  |  |  |
| High | - | 1·00 | - | - | 1·00 | - | - | 1·00 | - |
| Moderate | - | 1·31 (1·29-1·32) | - | - | 1·61 (1·57-1·65) | - | - | 1·20 (1·18-1·22) | - |
| Low | - | 1·35 (1·33-1·37) | - | - | 1·59 (1·55-1·63) | - | - | 1·30 (1·28-1·32) | - |
| Missing | - | 1·31 (1·29-1·33) | - | - | 1·08 (1·05-1·11) | - | - | 1·18 (1·16-1·21) | - |
| Healthcare degree |  |  |  |  |  |  |  |  |  |
| Yes | - | 1·00 | - | - | 1·00 | - | - | 1·00 | - |
| No | - | 1·38 (1·35-1·41) | - | - | 1·48 (1·43-1·54) | - | - | 1·39 (1·35-1·42) | - |

**Supplementary Table 5: Continued**

|  | **Flanders** | | | **Brussels** | | | **Wallonia** | | |
| --- | --- | --- | --- | --- | --- | --- | --- | --- | --- |
| **Variables** | **Adjusted OR (95%CI)** | | | **Adjusted OR (95% CI)** | | | **Adjusted OR (95%CI)** | | |
|  | **Model 1 ^a^** | **Model 2 ^b^** | **Model 3 ^c^** | **Model 1 ^a^** | **Model 2 ^b^** | **Model 3 ^c^** | **Model 1 ^a^** | **Model 2 ^b^** | **Model 3 ^c^** |
| Employment status |  |  |  |  |  |  |  |  |  |
| Employed | - | - | 1·00 | - | - | 1·00 | - | - | 1·00 |
| Unemployed | - | - | 1·56 (1·54-1·58) | - | - | 1·32 (1·30-1·35) | - | - | 1·40 (1·38-1·42) |
| Missing |  |  | 1·35 (1·31-1·39) | - | - | 0·94 (0·91-0·98) | - | - | 1·26 (1·20-1·32) |

^a^ Logistic regression model applied on all age groups (Flanders, n=3,348,547; Brussel, n=579,687; Wallonia, n=1,413,876). OR are adjusted for age, sex, region, migration background, household type, and income.

^b^ Logistic regression model applied only to individuals aged over 25 years (Flanders, n=2,955,807; Brussel, n=505,082; Wallonia, n=1,235,805). OR are adjusted for age, sex, region, migration background, household type, income, education level, and healthcare degree.

^c^ Logistic regression model applied only to individuals aged 25 to 65 years (Flanders, n=2,357,375; Brussel, n=436,356; Wallonia, n=998,369). OR are adjusted for age, sex, region, migration background, household type, and employment status. Income being strongly correlated with employment status, OR are not adjusted for income when employment status is included in the model.

#### Supplementary Table 6: Crude odds ratio (OR) and their 95% confidence interval (CI) for the association between sociodemographic and socioeconomic characteristics and the odds of not having received a first dose of COVID-19 vaccine, Belgium, December 28^th^ 2020 - August 31^st^ 2021.

| **Variables** | **Model 1 ^a^** | **Model 2 ^b^** | **Model 3 ^c^** |
| --- | --- | --- | --- |
|  | **Crude OR (95%CI)** | **Crude OR (95%CI)** | **Crude OR (95%CI)** |
| Age groups |  |  |  |
| 18-24 | 6·41 (6·26-6·56) |  | - |
| 25-34 | 6·68 (6·53-6·83) | 6·68 (6·53-6·83) | 3·13 (3·10-3·16) |
| 35-44 | 5·09 (4·98-5·21) | 5·09 (4·98-5·21) | 2·39 (2·36-2·41) |
| 45-54 | 3·28 (3·20-3·36) | 3·28 (3·20-3·36) | 1·54 (1·52-1·56) |
| 55-64 | 2·16 (2·12-2·21) | 2·16 (2·11-2·21) | 1·00 |
| 65-74 | 1·44 (1·40-1·48) | 1·14 (1·40-1·48) | - |
| 75-84 | 1·00 | 1·00 | - |
| 85+ | 1·09 (1·05-1·13) | 1·09 (1·05-1·13) | - |
| Sex ^a^ |  |  |  |
| Women | 1·00 | 1·00 | 1·00 |
| Men | 1·02 (1·01-1·02) | 0·99 (0·99-1·00) | 0·99 (0·98-0·99) |
| Region |  |  |  |
| Flemish | 1·00 | 1·00 | 1·00 |
| Brussels | 3·65 (3·62-3·68) | 3·52 (3·49-3·55) | 3·34 (3·31-3·37) |
| Walloon | 1·76 (1·75-1·77) | 1·81 (1·80-1·82) | 1·77 (1·75-1·78) |
| Migration background |  |  |  |
| Belgian natives | 1·00 | 1·00 | 1·00 |
| Second-generation migrants | 4·29 (4·25-4·33) | 3·70 (3·65-3·75) | 3·29 (3·25-3·34) |
| First-generation European migrants | 4·22 (4·19-4·26) | 3·98 (3·95-4·01) | 3·73 (3·70-3·76) |
| First-generation non-European migrants | 5·249 (5·211-5·286) ^*^ | 5·12 (5·08-5·16) | 4·52 (4·48-4·55) |
| Household type |  |  |  |
| Couples with children | 1·00 | 1·00 | 1·00 |
| Couples | 0·48 (0·48-0·49) | 0·47 (0·46-0·47) | 0·63 (0·62-0·63) |
| Single parents | 1·50 (1·48-1·51) | 1·48 (1·46-1·49) | 1·53 (1·51-1·54) |
| One-person households | 0·96 (0·95-0·96) | 0·94 (0·93-0·95) | 1·22 (1·21-1·23) |
| Collectivity | 0·29 (0·28-0·30) | 0·27 (0·26-0·28) | 0·62 (0·58-0·65) |
| Other | 1·50 (1·48-1·53) | 1·46 (1·44-1·49) | 1·59 (1·56-1·62) |
| Missing | 2·45 (2·38-2·52) | 2·31 (2·23-2·39) | 2·33 (2·25-2·41) |
| Income |  |  |  |
| High | 1·00 | 1·00 | - |
| Moderate | 1·74 (1·72-1·75) | 1·70 (1·68-1·72) | - |
| Low | 3·43 (3·40-3·46) | 3·12 (3·10-3·15) | - |
| Missing | 3·92 (3·87-3·97) | 3·35 (3·30-3·40) | - |
| Education level |  |  |  |
| High | - | 1·00 | - |
| Moderate | - | 1·65 (1·64-1·67) |  |
| Low | - | 1·56 (1·54-1·57) |  |
| Missing | - | 3·54 (3·51-3·57) |  |

**Supplementary Table 6: Continued**

| **Variables** | **Model 1^*^** | **Model 2^**^** | **Model 3^***^** |
| --- | --- | --- | --- |
|  | **Crude OR (95%CI)** | **Crude OR (95%CI)** | **Crude OR (95%CI)** |
| Healthcare degree |  |  |  |
| Yes | - | 1·00 | - |
| No | - | 1·71 (1·69-1·73) |  |
| Employment status |  |  |  |
| Employed | - | - | 1·00 |
| Unemployed | - |  | 1·67 (1·65-1·68) |
| Missing | - |  | 2·72 (2·68-2·77) |

^a^ Logistic regression model applied on all age groups (n=5,342,110).

^b^ Logistic regression model applied only to individuals aged over 25 years (n=4,696,694).

^c^ Logistic regression model applied only to individuals aged 25 to 65 years (n=3,792,100).

#### Supplementary Table 7*:* Crude odds ratio (OR) and their 95% confidence interval (CI) for the association between sociodemographic and socioeconomic characteristics and the odds of not having received a first dose of COVID-19 vaccine stratified by region, Belgium, December 28^th^ 2020 - August 31^st^ 2021.

|  | **Flanders** | | | **Brussels** | | | **Wallonia** | | |
| --- | --- | --- | --- | --- | --- | --- | --- | --- | --- |
| **Variables** | **Crude OR (95%CI)** | | | **Crude OR (95% CI)** | | | **Crude OR (95%CI)** | | |
|  | **Model 1 ^a^** | **Model 2 ^b^** | **Model 3 ^c^** | **Model 1 ^a^** | **Model 2 ^b^** | **Model 3 ^c^** | **Model 1 ^a^** | **Model 2 ^b^** | **Model 3 ^c^** |
| Age (years) |  |  |  |  |  |  |  |  |  |
| 18-24 | 6·90 (6·67-7·15) | - | - | 7·74 (7·32-8·19) | - | - | 4·31 (4·14-4·47 | - | - |
| 25-34 | 7·29 (7·04-7·54) | 7·29 (7·04-7·54) | 3·22 (3·17-3·27) | 5·38 (5·09-5·68) | 5·38 (5·09-5·68) | 2·42 (2·36-2·48) | 5·48 (5·28-5·69) | 5·48 (5·28-5·69) | 3·10 (3·04-3·15) |
| 35-44 | 5·52 (5·34-5·71) | 5·52 (5·33-5·71) | 2·44 (2·40-2·48) | 4·55 (4·31-4·81) | 4·55 (4·31-4·81) | 2·05 (2·00-2·10) | 4·01 (3·86-4·17) | 4·01 (3·86-4·16) | 2·27 (2·22-2·31) |
| 45-54 | 3·53 (3·41-3·66) | 3·53 (3·41-3·66) | 1·56 (1·54-1·59) | 3·16 (2·99-3·34) | 3·16 (2·99-3·34) | 1·42 (1·38-1·46) | 2·54 (2·45-2·64) | 2·54 (2·45-2·64) | 1·44 (1·41-1·47) |
| 55-64 | 2·29 (2·21-2·37) | 2·29 (2·21-2·37) | 1·00 | 2·25 (2·13-2·39) | 2·25 (2·13-2·39) | 1·00 | 1·79 (1·72-1·86) | 1·79 (1·72-1·86) | 1·00 |
| 65-74 | 1·49 (1·43-1·55) | 1·49 (1·43-1·55) | - | 1·47 (1·38-1·56) | 1·47 (1·38-1·56) | - | 1·26 (1·21-1·32) | 1·26 (1·21-1·32) | - |
| 75-84 | 1·00 | 1·00 | - | 1·00 | 1·00 | - | 1·00 | 1·00 | - |
| 85+ | 1·08 (1·02-1·14) | 1·08 (1·02-1·14) | - | 0·89 (0·81-0·98) | 0·89 (0·81-0·98) | - | 1·16 (1·10-1·23) | 1·16 (1·10-1·23) | - |
| Sex |  |  |  |  |  |  |  |  |  |
| Women | 1·00 | 1·00 | 1·00 | 1·00 | 1·00 | 1·00 | 1·00 | 1·00 | 1·00 |
| Men | 1·05 (1·04-1·06) | 1·02 (1·01-1·03) | 1·02 (1·01-1·03) | 0·96 (0·94-0·97) | 0·93 (0·92-0·94) | 0·91 (0·90-0·92) | 1·02 (1·01-1·03) | 1·01 (0·99-1·02) | 1·01 (0·99-1·02) |
| Migration background |  |  |  |  |  |  |  |  |  |
| Belgian natives | 1·00 | 1·00 | 1·00 | 1·00 | 1·00 | 1·00 | 1·00 | 1·00 | 1·00 |
| Second generation migrants | 4·55 (4·48-4·62) | 3·96 (3·88-4·04) | 3·51 (3·44-3·59) | 4·80 (4·68-4·93) | 3·26 (3·15-3·38) | 2·88 (2·78-2·99) | 2·59 (2·55-2·63) | 2·49 (2·44-2·54) | 2·22 (2·17-2·26) |
| First-generation European migrants | 5·63 (5·56-5·69) | 5·30 (5·23-5·37) | 5·02 (4·95-5·08) | 3·07 (3·01-3·14) | 2·83 (2·76-2·89) | 2·51 (2·45-2·58) | 2·18 (2·15-2·22) | 2·13 (2·10-2·16) | 2·03 (2·00-2·06) |
| First-generation non-European migrants | 5·54 (5·49-5·60) | 5·36 (5·30-5·42) | 4·64 (4·58-4·69) | 4·38 (4·29-4·47) | 4·25 (4·16-4·34) | 3·85 (3·77-3·94) | 3·40 (3·35-3·45) | 3·34 (3·29-3·39) | 2·95 (2·91-3·00) |

**Supplementary Table 7: continued**

|  | **Flanders** | | | **Brussels** | | | **Wallonia** | | |
| --- | --- | --- | --- | --- | --- | --- | --- | --- | --- |
| **Variables** | **Crude OR (95%CI)** | | | **Crude OR (95% CI)** | | | **Crude OR (95%CI)** | | |
|  | **Model 1 ^a^** | **Model 2 ^b^** | **Model 3 ^c^** | **Model 1 ^a^** | **Model 2 ^b^** | **Model 3 ^c^** | **Model 1 ^a^** | **Model 2 ^b^** | **Model 3 ^c^** |
| Household type |  |  |  |  |  |  |  |  |  |
| Couples with children | 1·00 | 1·00 | 1·00 | 1·00 | 1·00 | 1·00 | 1·00 | 1·00 | 1·00 |
| Couples | 0·49 (0·48-0·49) | 0·47 (0·46-0·48) | 0·64 (0·63-0·65) | 0·47 (0·47-0·48) | 0·49 (0·48-0·50) | 0·59 (0·58-0·61) | 0·56 (0·55-0·57) | 0·53 (0·52-0·54) | 0·71 (0·70-0·72) |
| Single parents | 1·52 (1·51-1·54) | 1·51 (1·49-1·53) | 1·56 (1·54-1·59) | 1·19 (1·17-1·21 | 1·18 (1·16-1·21) | 1·20 (1·18-1·23) | 1·38 (1·36-1·40) | 1·36 (1·33-1·38) | 1·40 (1·37-1·42) |
| One-person households | 0·95 (0·94-0·96) | 0·93 (0·92-0·94) | 1·24 (1·22-1·26) | 0·70 (0·69-0·71) | 0·72 (0·71-0·74) | 0·83 (0·81-0·85) | 0·97 (0·95-0·98) | 0·94 (0·93-0·96) | 1·21 (1·19-1·23) |
| Collectivity | 0·30 (0·29-0·32) | 0·28 (0·26-0·29) | 0·72 (0·67-0·78) | 0·23 (0·21-0·25) | 0·22 (0·20-0·24) | 0·45 (0·39-0·51) | 0·31 (0·29-0·33) | 0·29 (0·27-0·31) | 0·54 (0·49-0·59) |
| Other | 1·70 (1·66-1·74) | 1·66 (1·62-1·71) | 1·84 (1·79-1·89) | 0·83 (0·81-0·86) | 0·84 (0·81-0·87) | 0·84 (0·81-0·87) | 1·37 (1·32-1·41) | 1·30 (1·26-1·35) | 1·45 (1·39-1·50) |
| Missing | 2·98 (2·85-3·11) | 2·86 (2·72-3·00) | 2·89 (2·75-3·04) | 1·08 (1·03-1·14) | 1·02 (0·96-1·08) | 1·01 (0·95-1·07) | 2·20 (2·07-2·34) | 2·11 (1·97-2·27) | 2·15 (2·00-2·31) |
| Income |  |  |  |  |  |  |  |  |  |
| High | 1·00 | 1·00 | - | 1·00 | 1·00 | - | 1·00 | 1·00 | - |
| Moderate | 1·73 (1·71-1·76) | 1·69 (1·67-1·72) | - | 2·23 (2·18-2·29) | 2·15 (2·10-2·21) | - | 1·53 (1·51-1·55) | 1·50 (1·48-1·53) | - |
| Low | 3·33 (3·29-3·36) | 3·01 (2·98-3·05) | - | 3·62 (3·54-3·69) | 3·24 (3·17-3·32) | - | 2·47 (2·43-2·50) | 2·31 (2·28-2·35) | - |
| Missing | 4·54 (4·45-4·63) | 3·91 (3·82-3·99) | - | 2·94 (2·86-3·03) | 2·53 (2·46-2·61) | - | 2·30 (2·24-2·37) | 2·00 (1·93-2·06) | - |
| Education level |  |  |  |  |  |  |  |  |  |
| High |  | 1·00 | - | - | 1·00 | - | - | 1·00 | - |
| Moderate |  | 1·58 (1·56-1·60) | - | - | 2·48 (2·42-2·53) | - | - | 1·59 (1·57-1·62) | - |
| Low | - | 1·44 (1·42-1·46) | - | - | 2·25 (2·20-2·30) | - | - | 1·44 (1·42-1·47) | - |
| Missing | - | 4·31 (4·25-4·36) | - | - | 2·19 (2·15-2·24) | - | - | 2·42 (2·38-2·46) | - |

**Supplementary Table 7: continued**

|  | **Flanders** | | | **Brussels** | | | **Wallonia** | | |
| --- | --- | --- | --- | --- | --- | --- | --- | --- | --- |
| **Variables** | **Crude OR (95%CI)** | | | **Crude OR (95% CI)** | | | **Crude OR (95%CI)** | | |
|  | **Model 1 ^a^** | **Model 2 ^b^** | **Model 3 ^c^** | **Model 1 ^a^** | **Model 2 ^b^** | **Model 3 ^c^** | **Model 1 ^a^** | **Model 2 ^b^** | **Model 3 ^c^** |
| Healthcare degree |  |  |  |  |  |  |  |  |  |
| Yes | - | 1·00 | - | - | 1·00 | - | - | 1·00 | - |
| No | - | 1·73 (1·70-1·77) | - | - | 1·74 (1·68-1·80) | - | - | 1·45 (1·42-1·48) | - |
| Employment status |  |  |  |  |  |  |  |  |  |
| Employed | - | - | 1·00 | - | - | 1·00 | - | - | 1·00 |
| Unemployed | - | - | 1·70 (1·68-1·72) | - | - | 1·49 (1·47-1·51) | - | - | 1·33 (1·31-1·35) |
| Missing |  |  | 3·56 (3·47-3·65) | - | - | 1·26 (1·22_1·29) | - | - | 2·26 (2·17-2·35) |

^a^ Logistic regression model applied on all age groups (Flanders, n=3,348,547; Brussel, n=579,687; Wallonia, n=1,413,876).

^b^ Logistic regression model applied only to individuals aged over 25 years (Flanders, n=2,955,807; Brussel, n=505,082; Wallonia, n=1,235,805).

^c^ Logistic regression model applied only to individuals aged 25 to 65 years (Flanders, n=2,357,375; Brussel, n=436,356; Wallonia, n=998,369).
